# *De novo* truncation variants in the low-complexity C-terminal region of *XRN1* are associated with a dominant form of lethal infantile mitochondrial cardiomyopathy

**DOI:** 10.64898/2026.01.08.26343555

**Authors:** Liana N. Semcesen, Lucie S. Taylor, Leah E. Frajman, Marisa W. Friederich, Andrew M Frey, Stefan J. Siira, Daniella H. Hock, Tegan Stait, Sila Hopton, Yoshihito Kishita, Queenie K.-G. Tan, Vandana Shashi, Muriel Holder-Espinasse, Hugh Lemonde, Kay Metcalfe, Lisette Curnow, Rebecca C. Spillmann, Kelly Schoch, Karen Stals, Agata Oliwa, MitoMDT Diagnostic Network for Genomics and Omics, Network Undiagnosed Diseases, Matthias Trost, Kei Murayama, Yasushi Okazaki, Akira Ohtake, Aleksandra Filipovska, Charlotte L. Alston, John Christodoulou, David R. Thorburn, Johan L.K. Van Hove, Zornitza Stark, Robert W. Taylor, David A. Stroud, Alison G. Compton

## Abstract

*XRN1* encodes a highly conserved cytoplasmic 5’-3’ exoribonuclease involved in mRNA decay and quality control. It localizes to transient ribonucleoprotein aggregates, P-bodies and stress granules, where it interacts with other mRNA decay proteins and is involved in various cellular responses, including an emerging role in viral infection responses. Complete knockout of XRN1 in multicellular organisms is lethal, most likely due to its central role in mRNA homeostasis, with no prior human disease association reported. Here, we characterize seven individuals from six unrelated families with a lethal infantile form of mitochondrial cardiomyopathy caused by heterozygous *de novo* frameshift truncation variants clustering in the C-terminal region of *XRN1*, each predicted to evade nonsense-mediated mRNA decay. Each variant results in a near-identical XRN1 C-terminal sequence predicted to alter a characterized binding domain that interacts with the mRNA decapping enhancer EDC4. Biochemical investigations of striated muscle revealed combined oxidative phosphorylation deficiencies, demonstrated by decreased respiratory chain enzyme activities, decreased proteomics abundances, and abnormal histochemical reactivities. Despite having no established mitochondrial function in mammals, clinical and molecular findings across the cohort were consistent with mitochondrial disease. The precise mechanism by which the altered XRN1 proteins cause disease remains to be elucidated.

---

mRNA homeostasis is essential for cell health and survival by the process of ‘mRNA buffering’ which maintains a balance between mRNA synthesis and degradation. Several mRNA decay pathways exist in eukaryotes with most cytosolic mRNA processing occurring through the 5’-3’ exoribonuclease decay pathway.^1,2^

*XRN1* (MIM 607994, formerly known as *SEP1*) encodes the major eukaryotic and only known human cytosolic 5’-3’ exoribonuclease in this pathway, while its nuclear homolog *XRN2* (MIM 608851) is the analogous exoribonuclease within the nucleus. *XRN1* is highly conserved from humans (XRN1), to mice (Xrn1), flies (Pacman), worms (xrn-1) and yeast (Xrn1p).^1; 3^ In human cells, mRNAs destined for 5’-3’ decay undergo a highly coordinated degradation process. Initially, the multi-subunit CCR4-NOT complex catalyzes deadenylation to shorten the 3’ poly(A) mRNA tail. Subsequently, the DCP1/DCP2 decapping complex removes the protective 5’ m^7^G cap and the exposed mRNA body is rapidly degraded by XRN1 from the 5’ end.^2; 4; 5^ Proteins involved in the later stages of this pathway form networks of physical interactions facilitating the sequential handover of mRNA between the enzymatic steps, ensuring a tightly coupled and highly coordinated process. EDC4, a decapping enhancer, functions as a molecular scaffold that binds DCP1, DCP2, and XRN1, creating a localized processing hub. Additionally, XRN1 interacts with various subunits of the CCR4-NOT deadenylase complex and exhibits a weak association with the decapping activator PatL1.^6–9^

Beyond its well-established role in basal mRNA decay, XRN1 has been implicated in the processing and regulation of various RNA species, including selective turnover of tRNA species under heat stress conditions, and processing of rRNAs and long non-coding RNAs.^10;^11 Involvement of *Xrn1* in coordinating gene transcription, translation and decay has been demonstrated by interactions between Xrn1p/Xrn1 and the eukaryotic ribosome in both yeast and mouse models for mRNA surveillance and quality control,^5; 12^ and the role of Xrn1p in activating and directing mRNAs encoding membrane proteins to their translation site.^13^ More recently, XRN1 has been shown to participate in stress response mechanisms, contributing to stress granule (SG) dependent mRNA shortening during oxidative stress and regulating autophagy pathways under nutrient starvation conditions.^14; 15^ In the context of viral stress, XRN1 exhibits both antiviral and proviral properties depending on the specific pathogen.^16–18^

Loss of *Xrn1p* in *S. cerevisiae* cells causes severe growth defects and reduced viability,^19^ possibly due to absence of Xrn1p-Dcs1 (a scavenger decapping enzyme involved in degrading cleaved 5’-mRNA caps) activation which may contribute to growth defects through reduced levels of mitochondrial Por1 (homolog to human VDAC1).^20^ Depletion of Xrn1p in yeast has reported conflicting effects on global mRNA levels, depending on methodologies, with more recent studies showing an initial mRNA accumulation following Xrn1p depletion, followed by transcriptional compensation that returns mRNA levels to baseline through a feedback mechanism of mRNA abundance monitoring.^21^ Knockdown of *xrn-1* in *C. elegans* is embryonic lethal,^22^ as is whole-body knock-out in mice.^23^ However, a forebrain-specific *Xrn1* conditional knock-out mouse was investigated in the context of metabolic processing, with neuronal loss correlating to disrupted energy homeostasis.^23^ Knockdown in human cell lines also highlighted the role of XRN1 in mRNA buffering systems essential for adapting to environmental stimuli.^24^ While predominantly cytosolic, XRN1 concentrates in processing bodies (PBs), which are cytosolic ribonucleoprotein (RNP) foci that transiently form via liquid-liquid phase separation and contain mRNAs and mRNA decay machinery. Their precise role remains unresolved as P-body formation is not required for mRNA decay, although they likely serve as sites for coordinating and concentrating decay machinery.^7; 25^ PBs dynamically exchange RNA and protein components with stress granules (SGs), another species of phase-separated RNP condensates, with the composition of both adapting according to cellular needs. XRN1 has been found in both PBs and SGs under various cellular conditions, and both are interconnected in mRNA decay and stress responses.^7; 25; 26^ Recent work in *S. cerevisiae* suggests that Xrn1p hydrolyzes 5’ NAD-caps from a subset of mitochondrial mRNAs (mt-RNAs), releasing free NAD^+^ to regulate mitochondrial NAD^+^/NADH levels and contribute to mitochondrial homeostasis.^27^ While this study suggested human XRN1 can also hydrolyze NAD-capped mt-RNAs, it does so less efficiently than yeast Xrn1p, and does not consider differences in regulatory roles of 5’NAD caps between species.^28^ The mechanism by which cytosolic XRN1/Xrn1p acts on mt-RNAs is unclear, as no machinery for mt-RNA export from the mitochondrial matrix to the cytosol is known. Mitochondrial function and health are central to the pathogenicity of a wide range of diseases. Mitochondrial diseases represent a heterogenous group of genetic disorders caused by defects in mitochondrial DNA (mtDNA) or nuclear genes encoding mitochondrial proteins, often resulting in impaired oxidative phosphorylation (OXPHOS) and multi-system clinical manifestations.^29; 30^

In this work, we present evidence that supports heterozygous alterations in the C-terminus of *XRN1* as a cause of *de novo* dominant lethal infantile mitochondrial cardiomyopathy. In a collaborative effort facilitated via MatchMaker Exchange^31^ and personal communication, we have identified seven affected individuals from six unrelated families with similar clinical presentations of lethargy, poor feeding, lactic acidosis and hypertrophic cardiomyopathy, leading to cardiac failure and early death before one year of age (**Table 1**). All identified individuals were born to non-consanguineous parents with no family disease history of note. All individuals were born following uncomplicated pregnancies, with the exception of Family 3, where monozygotic twins (Individuals 3A and 3B) were delivered prematurely and experienced associated postnatal complications. Newborn hearing screens identified hearing impairment in 4 out of 7 individuals (**Table 1**). No other clinical abnormalities were observed at birth in any of the patients.

**Table 1.**
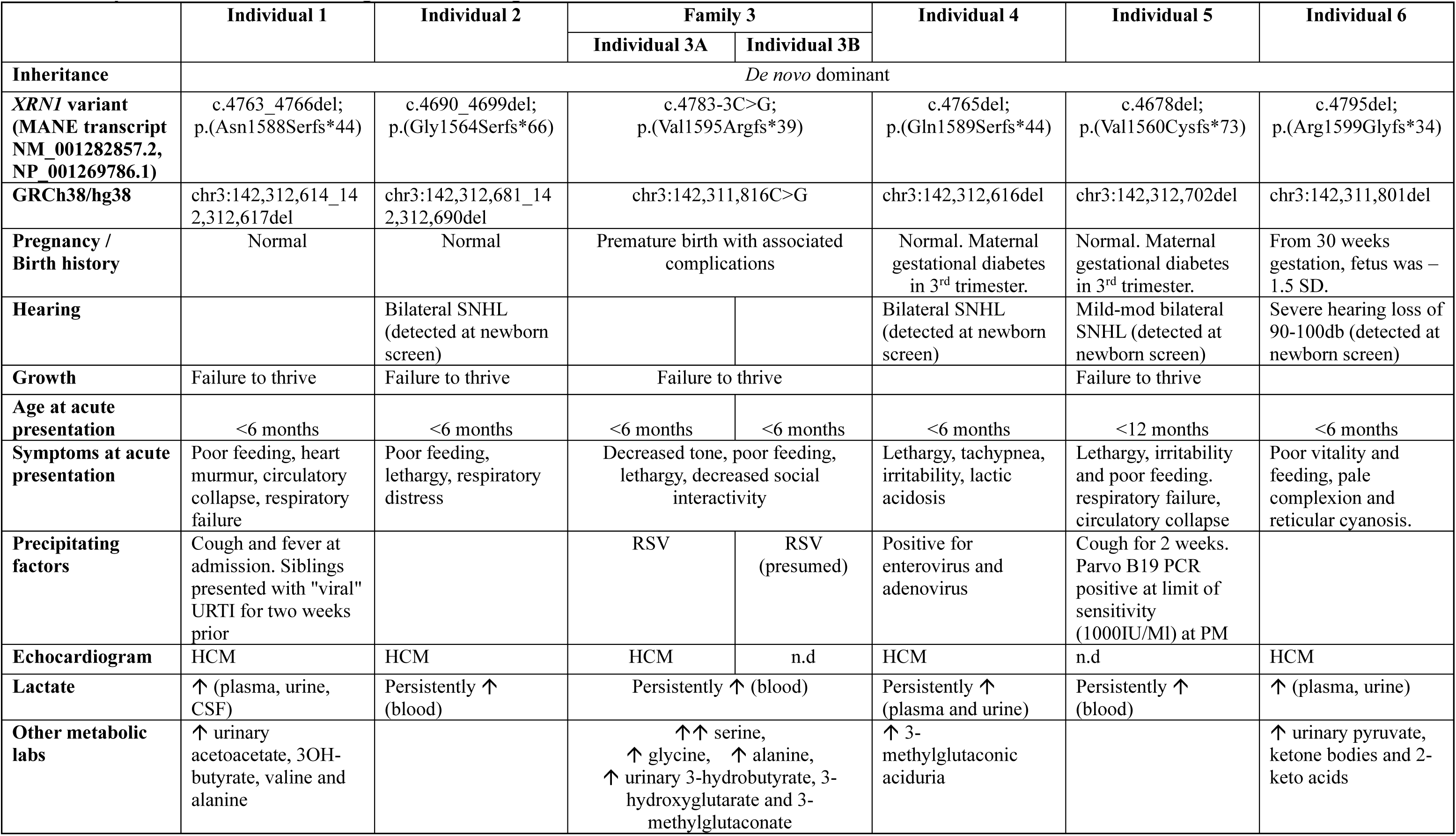
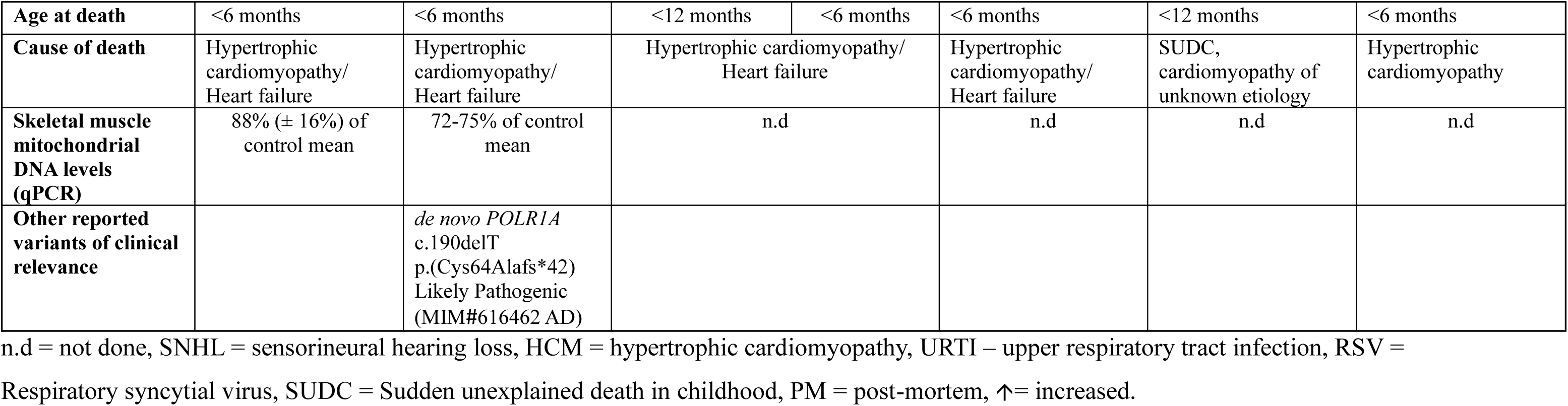
Key clinical, metabolic and genomic findings in individuals with *de novo* dominant *XRN1* C-terminal variants.

All individuals were admitted to hospital aged between 6 weeks and 11 months old following an acute onset of symptoms. Across the cohort, these symptoms were primarily characterized by poor feeding, lethargy, and irritability, and several of the individuals also reported hypotonia, failure to thrive, respiratory distress, and tachypnea. Symptoms were observed during or shortly after a suspected viral infection in most individuals. Individual 1 presented to hospital with a fever and a cough two weeks after their siblings experienced a suspected viral upper respiratory tract infection. Individuals 3A and 3B were both admitted to hospital with respiratory syncytial virus (RSV). Individual 4 tested positive for adenovirus and enterovirus during hospitalization. Individual 5 had a cough which resolved prior to hospital admission. Two weeks earlier, both their parents had a mild SARS-CoV-2 infection, and the patient tested PCR-positive to Parvovirus B19 in postmortem analysis. No viral episodes were noted in Individuals 2 and 6. Elevated lactate levels were measured in either blood, plasma or urine in all individuals. Multiple urinary metabolites were elevated across the cohort. Both individuals from Family 3 showed increased levels of 3-hydroxyglutarate and 3-methylglutaconate. Individual 4 had increased 3-methylglutaconic aciduria, while Individual 6 had increased urinary pyruvate, ketone bodies and 2-keto acids. Hypertrophic cardiomyopathy was confirmed by echocardiogram in all individuals except for Individuals 3B and 5, for whom the test was not performed. However, all affected individuals were confirmed to have died from hypertrophic cardiomyopathy and heart failure on autopsy (**Table 1**). Detailed clinical descriptions and genetic histories are described in **Supplemental notes: Case Reports**.

Clinical features of all individuals in the cohort were suggestive of mitochondrial disease, prompting respiratory chain enzymology to measure the activity of mitochondrial complexes I-IV. These results indicated a mitochondrial respiratory chain defect in all individuals tested (**Table 2**). Individual 1 exhibited severe loss of skeletal muscle Complex I, III and IV activities (8%, 19%, and 11% of control means, respectively). Activities of Complexes I and IV were also severely lost in cardiac muscle (2% and 23%, respectively), while liver enzymology showed borderline low activities for Complexes I and IV (37% and 53%, respectively) and activities in subject-derived skin fibroblasts were largely unremarkable. Individual 2 demonstrated markedly decreased skeletal muscle activities for Complexes I, III and IV (11%, 25% and 4%, respectively) and borderline low for Complex IV activity in liver (31%), while fibroblast enzymology was normal. Skeletal muscle enzymology of Individuals 3A and 3B both revealed a severe loss of Complex I activity (13% and 6%, respectively), while Complex IV activity was mildly decreased in Individual 3A (44%) and markedly decreased in Individual 3B (18%). Individual 4 showed severe combined skeletal muscle deficiencies of Complexes I and IV with normal fibroblast enzymology. Individual 5 skeletal muscle revealed a significant deficiency in Complex I and IV activities (4% and 7%, respectively). Similar deficiencies were observed in cardiac muscle of Individual 5 (4% and 29%, respectively), and fibroblast enzymology remained within normal limits for both individuals. No autopsy samples were available for Individual 6 to facilitate either respiratory chain enzymology or other functional studies. Quantitative real-time PCR to investigate mtDNA depletion was performed on skeletal muscle DNA for Individuals 1 and 2 and was within normal ranges (**Table 1**).

**Table 2.** Respiratory Chain Enzymology in Tissues and Cells.

| Individual # | Tissue | Respiratory Chain Enzymology (% Activity) |  |  |  |  |  |
| --- | --- | --- | --- | --- | --- | --- | --- |
|  |  | CI | CII | CII+III | CIII | CIV | CS |
| Individual 1 | Skeletal Muscle | **8** | 318 | 72 | **19** | **11** | 207 |
|  | Cardiac Muscle | **2** | 75 | **25** | 123 | **23** | 121 |
|  | Fibroblasts | 28 | 72 | 77 | 25 | 33 | 50 |
|  | Liver | 37 | 82 | n.d | 84 | 53 | 69 |
| Individual 2 | Skeletal Muscle | **11** | 443 | 58 | **25** | **4** | 272 |
|  | Fibroblasts | 142 | 236 | 94 | 230 | 107 | 106 |
|  | Liver | 121 | 155 | n.d | 153 | 31 | 111 |
| Individual 3A | Skeletal Muscle | **13** | 109 | 82 | 48 | 44 | 116 |
| Individual 3B | Skeletal Muscle | **6** | 189 | 62 | 47 | **18** | 155 |
| Individual 4 | Skeletal Muscle <sup>a</sup> | **0.011**<br>(0.118-0.332) | n.d | **0.009**<br>(0.072-0.335) | n.d | **not detected**<br>(0.013-0.039) | n.d |
|  | Fibroblasts | 102 | 203 | n.d | 103 | 143 | n.d |
| Individual 5 | Skeletal Muscle | **4** | 60 | n.d | 94 | **7** | n.d |
|  | Cardiac Muscle | **4** | 66 | n.d | 50 | **29** | n.d |
|  | Fibroblasts | 92 | 170 | n.d | 188 | 172 | n.d |

CI= NADH:Coenzyme Q<sub>1</sub> oxidoreductase; CII= succinate:Coenzyme Q<sub>1</sub> oxidoreductase; CII+III= succinate:cytochrome *c* oxidoreductase; CIII= = decylbenzylquinol:cytochrome *c* oxidoreductase; CIV= cytochrome *c* oxidase; CS= citrate synthase; n.d = not done; Deficient values are marked in bold; No samples were available for Individual 6; % activities expressed relative to protein as % of control mean activity. <sup>a</sup> RCE for Individual 4 Skeletal Muscle is represented as values relative to the upper and lower limits of the control range, rather than % activity.

Clinical genome and/or exome sequencing did not identify any mitochondrial disease related variants that could account for the observed phenotypes, although independently undertaken data reanalysis identified distinct *de novo* heterozygous variants affecting the last two exons of *XRN1* (NM_001282857.2) (**Figure 1A**), each resulting in a shared C-terminal protein sequence and premature truncation (**Figure 1B**). Individual 1 carries a c.4763_4766del; p.(Asn1588Serfs*44) variant and Individual 2 a c.4690_4699del; p.(Gly1564Serfs*66) variant, both predicted to escape nonsense-mediated mRNA decay (NMD). Family 3 (Individual 3A and 3B) were found to have a heterozygous *de novo* splice variant in *XRN1* c.4783-3C>G, predicted to abolish the canonical acceptor site of the final exon (SpliceAI score^32^ 0.91 and Pangolin score^33^ 0.84), and result in protein truncation at p.(Val1595Argfs*39) and the creation of the same common C-terminal motif. Individual 4 has a heterozygous *de novo* c.4765del; p.(Gln1589Serfs*44) variant and Individual 5 a heterozygous *de novo* c.4678del; p.(Val1560Cysfs*73) variant. Finally, Individual 6 was found to have a heterozygous *de novo* c.4795del; p.(Arg1599Glyfs*34) variant. All variants are predicted to create the same C-terminal truncating motif across the cohort and were confirmed to be *de novo* through trio genome sequencing (GS) or targeted sequencing of parental samples. Each *XRN1* variant was absent from the population database gnomAD v4.1.0,^34^ excluding Individual 6 whose variant was detected in heterozygosity in 5 exome samples within the gnomAD v4 set. All 5 samples had an allelic balance between 0.2-0.25 and low depth of coverage with 4 at 10-15x and 1 at 20-25x, whereas in Individual 6 it was detected in 24 of 40 reads by GS, raising the possibility that the gnomAD individuals could be mosaic for this variant. *XRN1* has a pLI^35^ of 1, a pLOEUF score^36^ of 0.298, a sHet score^37^ of 0.047 and a pHaplo score^37^ of 0.92, all of which suggest that this gene is likely to be intolerant of variants that would result in a change in dosage. cDNA studies using RNA extracted from fibroblasts from Individuals 1, 2 and skeletal muscle obtained from Individual 4 (**Figure S1**) confirmed that both wild-type and mutant alleles were present, supporting the prediction that the mutant alleles would escape NMD.

**Figure 1:**
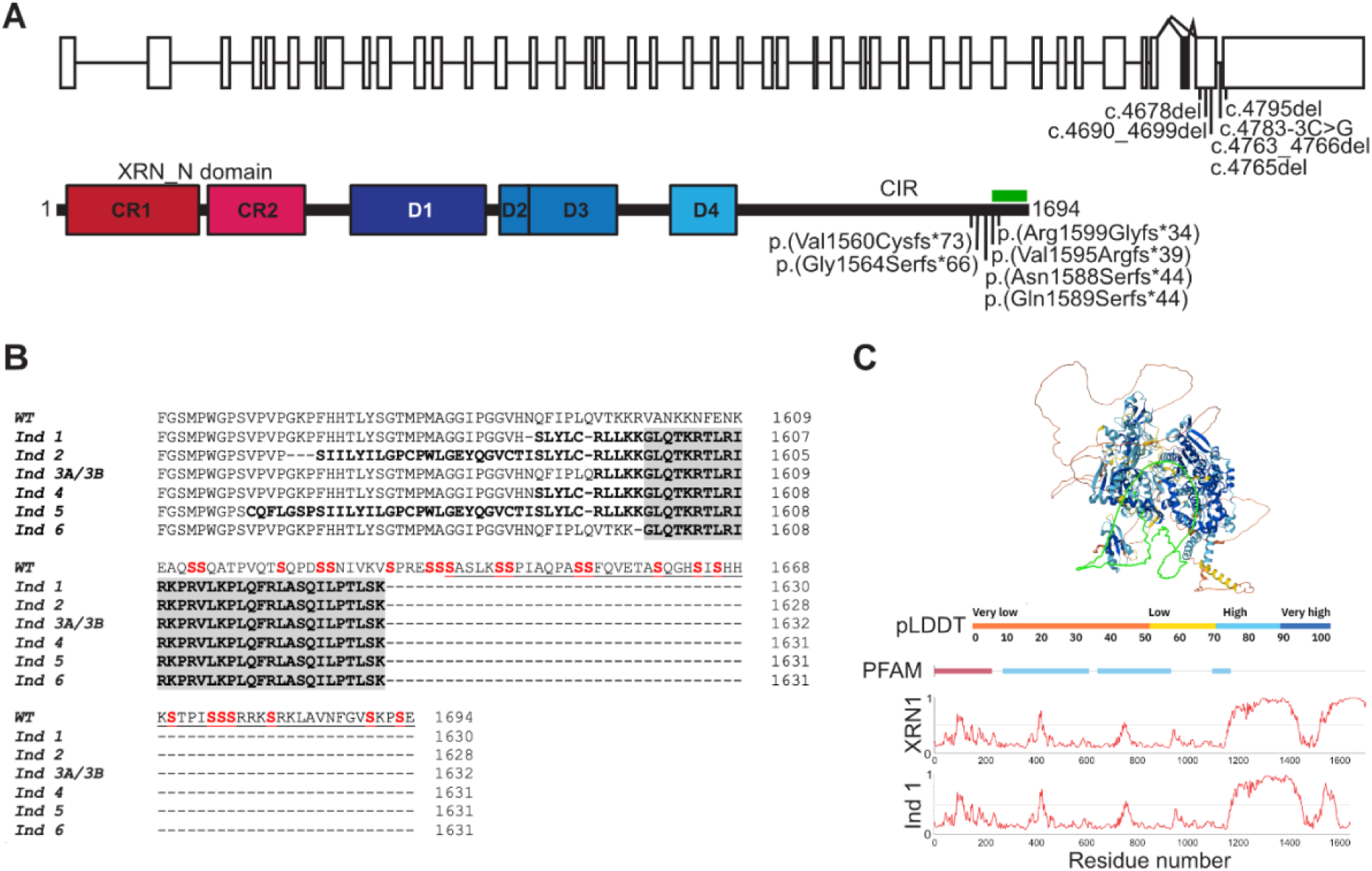
De novo C-terminal variants in XRN1 result in an identical truncating protein motif which alters the low-complexity region and EDC4-binding domain. (A) Exon structure of the XRN1 transcripts (NM_001282857.2 and NM_019001.5) which differ by an additional in-frame exon (shown filled in black) in NM_019001.5. NM_001282857.2 is the MANE transcript and the predominant one detected in the 54 tissue types within the GTEx dataset including skeletal muscle, heart and fibroblasts.^55^ The six different variants identified in this study are annotated to the MANE transcript. XRN1 protein (NP_001269786.1) contains two highly conserved regions within its N-terminal XRN domain (shaded in red tones, CR1 and 2) which has catalytic activity, along with 4 conserved domains (D1-D4 in blue)^38^ that are thought to stabilize the N-terminal domain and a low-complexity region comprising the C-terminal interacting region (CIR) containing the EDC4-binding motif (green bar).^8; 9^ (B) Sequence alignment of C-terminal XRN1 amino acids 1551 to 1694 of affected individuals (Ind 1 - 6) compared to wild-type (WT) demonstrating that the truncating variants identified in this study all result in the loss of the serine-rich region (red) along with the EDC4-binding region (underlined in WT) and create an identical conserved motif (grey). Bold black are the mutated residues. (C) The AlphaFold structure prediction,^39^ generated August 1^st^ 2025, (top) of human XRN1 and the predicted disorder tendency (bottom), illustrate substantial disorder in the C-terminus (after position 1200) which is altered in these affected individuals (Ind 1 illustrated in this figure compared to WT). This region is highlighted in green in the AlphaFold structure and the prediction profile is generated using UniProt: Q8IZH2 the default options (AIUPred^56^ - only disorder, Default smoothing). The AlphaFold structure is colored as per its calculated pLDDT (predicted local distance difference test) score, representing the confidence of the predicted structure. Annotated regions from PFAM (PFAM row), red is XRN_N domain and blue are D1, D2/3 and D4 domains colored as per part (A).

The C-terminal end of human XRN1 contains a low-complexity region^9^ that has been shown to be necessary for its binding to EDC4.^6–8^ This C-terminal region is absent in yeast Xrn1p, and while the crystal structure of *Drosophila* Pacman (Xrn1) was recently elucidated, it did not include this unstructured region.^38^ This region is also ill-defined in AlphaFold models^39; 40^ with a per-residue model confidence score (pLDDT) of 69.62. Alignment of wildtype XRN1 with the predicted patient proteins showed that along with the loss of the 56 most C-terminal residues (underlined in **Figure 1B**) necessary for EDC4 binding, all patient variants led to the creation of a shared common protein motif (**Figure 1B**). This truncation alters the serine composition within the C-terminus, where the enrichment of polar amino acids is known to contribute to disordered regions of proteins and drives protein and RNA interactions.^41^ The resulting C-terminal sequence exhibits an altered amount of disorder in the C-terminus of XRN1 (**Figure 1C**).

Despite *XRN1* having no known function relating to mitochondria in mammals, the striking similarity in clinical presentation and shared C-terminal motif suggests that *XRN1* is the disease gene in this cohort. We performed mass spectrometry based untargeted quantitative proteomics on whole-cell skeletal muscle lysates from Individuals 1-5 to better understand the extent of the mitochondrial phenotype. Proteomics based Relative Complex Abundance (RCA) analysis, which has previously been benchmarked against respiratory chain enzymology for diagnosing mitochondrial disorders,^42^ revealed a shared combined OXPHOS defect predominantly affecting the overall abundances of Complexes I, III and IV, consistent with enzymology findings. Skeletal muscle from Individual 1 showed RCA deficiencies in Complexes I, III and IV (21%, 44% and 17% of control means, respectively), while Individual 2 showed a similar pattern of reduced abundances (20%, 47% and 12% respectively) (**Figures 2A and 2B**). Similarly, Individuals 3A, 3B and 4 demonstrated reduced Complex I and IV abundances (3A: 33% and 35%; 3B: 21% and 20%; 4: 18% and 13%, respectively), with Complex III abundance also decreased in these individuals, although not reaching the statistical significance observed in Individuals 1 and 2 (**Figures 2C, 2D and 2E**). Individual 5 RCA analysis measured decreased abundances of Complexes I, III and IV (40%, 72% and 29%, respectively) (**Figure 2F**). While RCA profiles were largely consistent with enzymology results (**Table 2**), RCA analysis demonstrated increased sensitivity, detecting reduced abundance of Complex IV in Individual 3A and Complex III in Individual 5 that were not observed in enzymology of the same sample. Individual 3A RCA analysis also detected a unique reduction in the abundance of Complex V (77% of control mean) (**Figure 2C**), though Complex V levels have been shown to have greater variability in overall abundance.^42^

**Figure 2:**
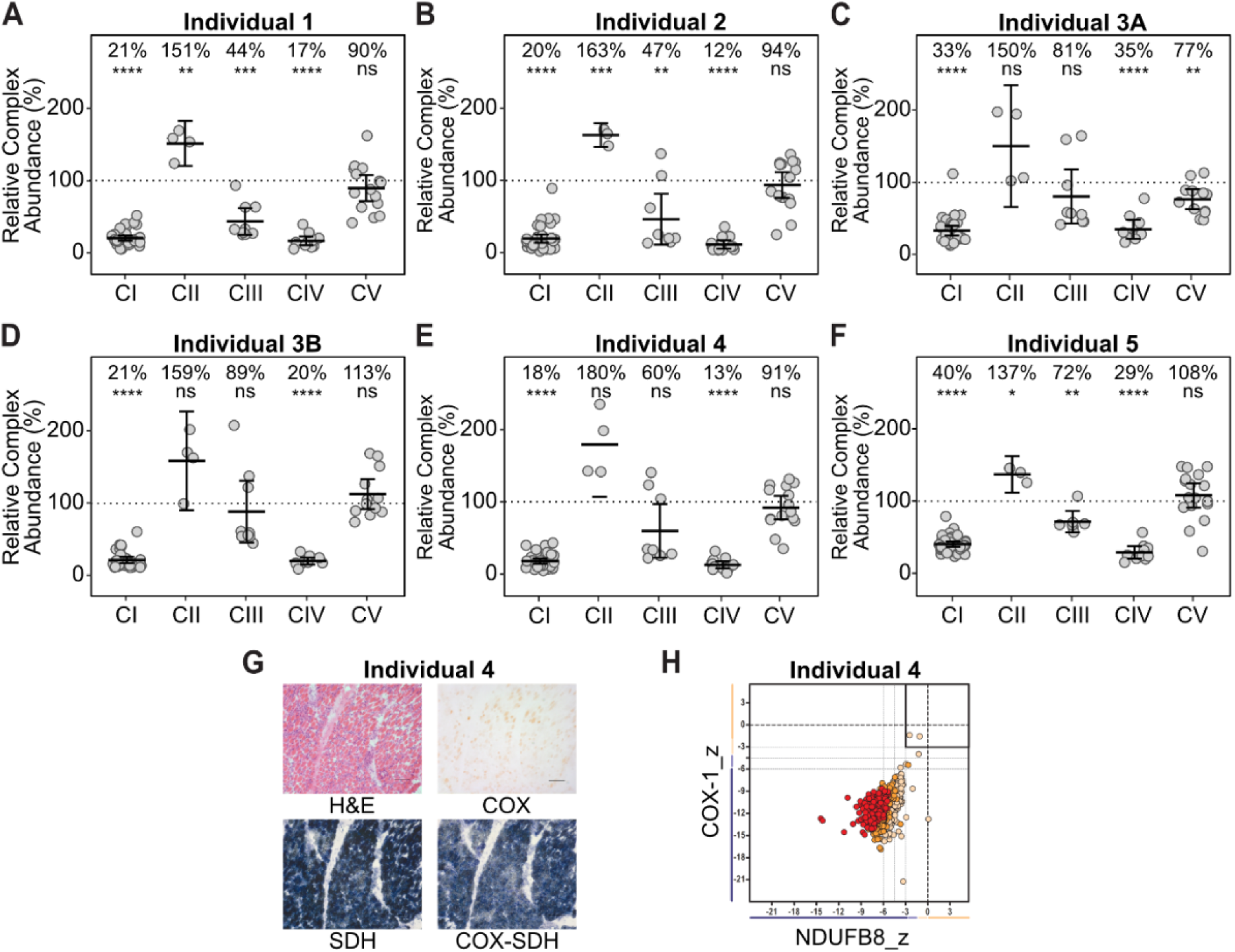
Quantitative proteomics and immunohistochemistry support the biochemical diagnosis of a mitochondrial disorder manifesting as a combined OXPHOS defect inskeletal muscle. (A-F) Relative Complex Abundance (RCA) of OXPHOS Complexes I-V (CI-V) from skeletal muscle proteomic data of each indicated individual compared to controls (n=3). Individual protein subunits are represented as a single dot and the mean value of each complex is indicated by the middle bar and represented as a percentage. Upper and lower bars represent 95% confidence intervals. Significance was calculated from a two-sided t-test. ns = not significant, * = p<0.05, ** = p≤0.01, *** = p≤0.001, **** = p≤0.0001. (G) Histopathological assessment of frozen muscle cryosections from Individual 4 revealed significant abnormalities. Hematoxylin and eosin (H&E) staining shows general muscle morphology. Cytochrome c oxidase (COX) and sequential succinate dehydrogenase (SDH) histochemistry reveal a global loss of COX enzyme activities, whilst SDH histochemical activity is strongly preserved, in agreement with the proteomic data. Scale bar = 100 µm. (H) Quadruple immunofluorescence analysis of NDUFB8 (Complex I) and COXI (Complex IV) expression in skeletal muscle from Individual 4. Each dot represents the measurement from an individual muscle fiber, color coordinated according to its mitochondrial mass (low = blue, normal = beige, high = orange, very high = red). Gray dashed lines represent SD limits for classification of the fibers. Lines next to x- and y- axis represent the levels of NDUFB8 and COXI: beige = normal (>-3), light beige = intermediate positive (-3 to -4.5), light purple = intermediate negative (-4.5 to -6), purple = deficient (<-6). Bold dashed lines represent the mean expression level of normal fibers. These data confirm a loss of both NDUFB8 and COX1 expression, consistent with the multiple OXPHOS defects demonstrated by enzymology and proteomics.

Further functional studies confirmed combined OXPHOS deficiencies in skeletal muscle. Blue Native PAGE (BN-PAGE) with in-gel activity assays of muscle mitochondrial fractions showed decreased Complex I and IV assembly and activities, and an accumulation of a lower molecular weight Complex V band in Individuals 3A and 3B, suggestive of a combined deficiency due to an impairment in the processing of mtDNA encoded subunits (**Figures S2A and S2B**). BN-PAGE analysis of both Individual 4 and 5 skeletal muscle similarly showed diminished assembly of Complexes I, IV, and V (**Figures S2C and S2D**). Furthermore, skeletal muscle histochemistry from Individuals 4 and 5 indicated coarse SDH reactivity and absent COX (Complex IV) reactivity, reminiscent of the histopathological mitochondrial dysfunction seen in many mitochondrial disease patients (**Figures 2G and S2E**). This was corroborated by quadruple OXPHOS immunofluorescence staining showing marked loss of MT-CO1 (Complex IV) and NDUFB8 (Complex I) immunoreactivity in Individual 4 (**Figure 2H**). While similar combined OXPHOS deficient profiles were not seen by RCA analysis of fibroblasts from Individuals 1, 2 and 4 (**Figures S3A, 3B and 3C**), they were observed by RCA analysis, BN-PAGE and histochemistry of cardiac muscle from Individual 1 and 5 (**Figure S2D, S2F, S3D-E**), consistent with the respiratory chain enzymology results (**Table 2**). The clinical presentation of hypertrophic cardiomyopathy accompanied by the functional OXPHOS deficiency in skeletal and cardiac muscle indicates an organ-specific clinical presentation in these patients affecting primarily striated muscle tissues. Diseases affecting mitochondrial functions typically present with multi-systemic phenotypes, however isolated mitochondrial cardiomyopathy has been reported in several cases, not uncommonly in disorders of mitochondrial RNA processing and translation disorders.^30; 43^ The underlying mechanism for tissue-specific presentations in mitochondrial disorders remains poorly understood. In this cohort, the onset of symptoms was rapid following viral infections in most individuals, and the high energy requirements of the heart make it particularly vulnerable to oxidative stress and mitochondrial disorders. The consistent and specific pattern of OXPHOS deficiencies across individuals strongly suggests a shared underlying mechanism, reinforcing the pathogenicity of the *XRN1* variants despite the unknown link to mitochondrial functioning.

To elucidate how these *XRN1* variants could cause the observed mitochondrial dysfunction, we first assessed the similarity of the OXPHOS presentations detected by proteomics (**Figure 3A**). A heatmap of the log2 fold-change values for OXPHOS complex subunits from the skeletal muscle proteomics emphasized the high degree of similarity in OXPHOS presentation at a subunit level, indicating each of the C-terminal *XRN1* variants similarly impairs mitochondrial function. To investigate whether the mitochondrial dysfunction observed in these patients results from defective processing of mt-RNAs by XRN1 in relation to the suggested NAD de-capping role of yeast Xrn1p,^27^ we analyzed mt-RNA processing and expression by RNA-sequencing (RNAseq). Analysis of skeletal muscle obtained from Individuals 1, 2 and 4 revealed no significant deficiencies in mt-RNA expression levels compared to controls (**Figure S4A**). Only a small percentage of the human mt-RNA population is predicted to harbor a non-canonical 5’ NAD cap,^44^ and the mechanism by which cytosolic XRN1 could access mt-RNAs is not clear, as both the wildtype and the mutant XRN1 sequences we report in this study have no predicted mitochondrial localization signal. Additionally, mitochondrial DNA and RNA processing pathways differ significantly between humans and yeast, with yeast mtDNA and mt-RNA requiring considerably more extensive processing compared to human.^45^ The role of human XRN1 in mt-RNA processing requires further investigation, although we do not observe a decrease in mt-RNA as the cause of the mitochondrial phenotypes in this cohort. Levels of *XRN1* transcripts were also unchanged between patients and controls. RNAseq of skeletal muscle additionally showed elevated ADP/ATP translocase 2 (*SLC25A5* MIM 300150) transcripts as a possible compensatory response to imbalanced OXPHOS levels. In addition, the expression of stress response markers mitokine *GDF15* (MIM 605312) and *ATF3* (MIM 603148) were elevated (**Figure S4A**), consistent with cardiomuscular-manifesting pathologies previously identified in mitochondrial diseases and the mitochondrial disease profiles of this cohort.^46–49^

**Figure 3:**
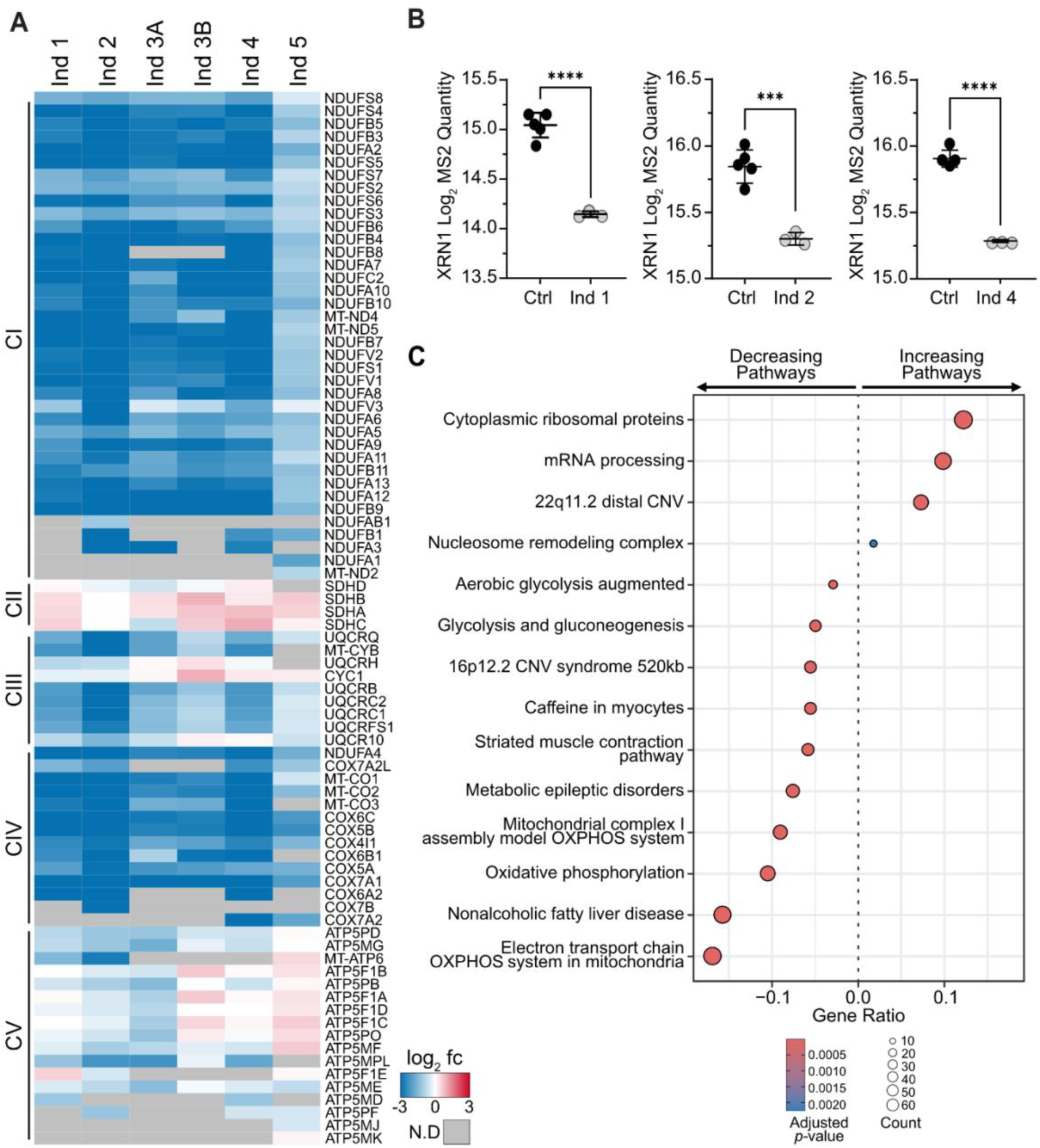
Proteomic profiling reveals consistent mitochondrial dysregulation, altered XRN1 protein levels and altered cellular pathways. (A) Heatmap depicting log2 fold-changes of OXPHOS subunits in skeletal muscle proteomics data. Each individual is compared against controls from the same experimental batch. N.D = not detected. **(B)** Relative abundance of XRN1 protein in fibroblast proteomics from Individual 1 (left), Individual 2 (middle) or Individual 4 (right) compared to controls. Significance was calculated from an unpaired t-test between the means. Error bars indicate standard deviation. *** = p≤0.001, **** = p≤0.0001. **(C)** Enrichment analysis of genes significantly increased or decreased (p≤0.05, fold-change ± 1.5) in abundance in muscle proteomic data of at least two of the following probands: Individuals 1, 2,4 and 5. Individuals 3A and 3B were excluded due to lower detection of non-mitochondrial proteins. Pathways were annotated using WikiPathways and determined within R using the clusterProfiler package.

Proteomics analysis of available fibroblast cell lines (Individuals 1, 2 and 4) identified a significant decrease in XRN1 protein abundance in each individual when compared with controls (**Figure 3B**) and was confirmed by immunoblot analysis (**Figure S4B**). Although ubiquitously expressed across tissue types, XRN1 was unable to be quantified by either technique in skeletal or cardiac muscle of both control and patient samples, due to technical limitations. We were unable to distinguish between expression of the wildtype or mutant alleles in quantified XRN1 protein in fibroblasts, as no peptides unique to the wildtype or altered C-terminal region were detected in either control or patient samples. Moreover, the antibody used by immunoblot analysis is specific for only the shared regions of the wildtype and mutant protein. Although we confirmed that the mutant *XRN1* transcripts are stably expressed in relevant tissues and predicted to escape NMD by both RNAseq and cDNA studies, we have not been able to assess the stability of the resulting mutant protein, and therefore cannot differentiate between a loss-of-function disease mechanism due to haploinsufficiency from one *XRN1* allele, or a gain-of-function disease mechanism due to the presence of the potentially toxic C-terminal XRN1 sequence. Notably, it has been shown that complete loss of XRN1 expression is embryonically lethal in multi-cellular organisms,^19; 22^ which may explain the heterozygosity of these individuals and suggests that a gain-of-function mechanism may be more likely than complete loss of function.

The C-terminal XRN1 interactor EDC4 was only detected in skeletal muscle proteomic datasets of Individuals 2, 4 and 5, where it was significantly increased in abundance relative to controls (**Figure S4C**). Fibroblast proteomics saw varying levels of EDC4 across individuals, suggesting the abundance of EDC4 is not dependent on the presence or interaction with XRN1 C-terminal EDC4 binding domain (**Figure S4D**). Other mRNA decay proteins were inconsistently detected in skeletal muscle samples and although detected in fibroblasts samples, showed no clear pattern of differential expression (**Figure S4D**). We performed enrichment analysis on proteins with significantly increased or decreased abundances (*p* ≤ 0.05, fold-change ± 1.5) in the skeletal muscle proteomics data of at least two affected individuals, highlighting terms relating to mitochondrial energy production as the most significantly downregulated across the cohort (**Figure 3C**). In keeping with the known functions of XRN1, such as mRNA decay and translational repression, pathways associated with cytosolic ribosomal proteins and mRNA processing were consistently upregulated across the cohort, although genes associated within the enriched ‘mRNA processing’ term are largely involved in pre-mRNA splicing rather than mature mRNA decay. Pathways associated with both 22q11.2 and 16p12.2 copy number variants (CNV) were also significantly enriched; however, genes annotated to these pathways largely overlap with ‘OXPHOS’, ‘cytoplasmic ribosomal proteins’ and ‘mRNA processing’ terms, reflecting an overrepresentation of these functional pathways rather than CNV syndromes in these individuals (**Table S5**).

Many roles have been attributed to *XRN1,* with extensive investigations conducted using its yeast ortholog, *Xrn1p*. While it is currently unclear which of these functions account for the mitochondrial phenotype seen in this cohort, the involvement of *XRN1* in both mammalian mRNA decay and viral stress responses may be implicated, although the disease mechanism is not explored in depth in this study. The different C-terminal variants in these individuals are all predicted to alter interactions between XRN1 and EDC4. Disruptions in XRN1-EDC4 binding and stoichiometry have shown to directly impact mRNA decay rates, leading to larger sized PBs which in turn interferes with mRNA decapping and suppresses mRNA expression.^7^ Specifically, PB formation has been shown to support cellular viability and mitigates SG formation when XRN1 levels are decreased.^7^ These identified frameshift variants alter the intrinsically disordered C-terminal region of XRN1 and may interfere with its ability to localize into PBs or SGs, as frameshift variants in disordered regions have been implicated in altered phase separation of biochemical condensate dynamics contributing to disease pathogenicity.^50; 51^ Beyond mRNA, recent studies have demonstrated that XRN1 is essential for proper innate immune activation in viral mimicking conditions.^52; 53^ Loss of XRN1 in these viral conditions led to accumulation of cytoplasmic dsDNA and reduced cell viability, triggering activation of immune receptors MDA5 and PKR, and downstream mitochondrial and peroxisomal anti-viral signaling protein MAVS. Notably, most individuals within this cohort experienced a viral infection prior to onset of symptoms. The presence of the heterozygous *de novo* C-terminal variant or decrease in XRN1 abundance may impact cellular responses to viral infections, as has been shown in the cancer cell lines.^52; 53^ Additionally, viral infections can induce SG, and in turn PB formation.^54^ Disruption of either or a combination of these roles, antiviral responses or PB dynamics, may contribute to the observed phenotypes in these individuals, however, the precise mechanism of linking XRN1 dysfunction to the mitochondrial defects remains unclear.

Taken together, our clinical, genetic and biochemical studies define an autosomal dominant disorder caused by *de novo* heterozygous variants altering the last two exons of *XRN1* leading to an identical unique C-terminal motif presenting as a lethal infantile form of mitochondrial hypertrophic cardiomyopathy. A consistent and isolated oxidative phosphorylation phenotype affecting post-mitotic striated (skeletal and cardiac) muscle tissues was observed in these individuals and are reminiscent of a perturbation in the synthesis of mtDNA encoded subunits through impact of their processing. However, the disease mechanism and precise pathway in which defective XRN1 leads to mitochondrial dysfunction and associated disease phenotype requires further investigation.

## Declaration of interests

The authors declare no competing interests.

## Consortia

### MitoMDT Diagnostic Network for Genomics and Omics

David R Thorburn, Aleksandra Filipovska, Michael T Ryan, David A Stroud, Diana Stojanovski, David Coman, Sean Murray, Ryan L Davis, John Christodoulou, Suzanne CEH Sallevelt, Roula Ghaoui, Cas Simons, Stefan J Siira, Shanti Balasubramaniam, Alison G Compton, Daniel G MacArthur, Nicole J Lake, Drago Bratkovic, Joy Lee, Maina Kava, Amanda Samarasinghe, Yoni Elbaum, Catherine Atthow, Pauline McGrath, Tegan Stait, Rocio Rius, Liana Semcesen, Megan Ball, Daniella Hock, Luke Formosa, Ellenore M Martin, Madeleine Harris, Leah Frajman.

### Undiagnosed Diseases Network (UDN)

Authorship list provided in supplemental notes

## Data Availability

The raw genomic and proteomic data that supports the findings of this study have not been deposited in a public repository due to human research ethics committee (HREC) and institutional review board (IRB) restrictions but are available from the corresponding authors on request. Variants described here were deposited in ClinVar before publication.

## Acknowledgments

Acknowledgments and funding sources provided in supplemental notes.

## Author contributions

A.G.C, D.A.S, R.W.T, C.L.A, J.L.K.V.H, D.R.T, J.C conceived and supervised the study; Z.S, Q.K.G.T, M.H-E, Ka.M, Ke.M, Ak.O, H.L, V.S, L.C, R.C.S, Ke.S recruited patients and/or assessed the clinical information; A.G.C, C.L.A, Ka.S, Ag.O, Y.K, Y.O analyzed the genomic data; M.W.F, T.S performed respiratory chain analyses; L.N.S, L.S.T, L.E.F, M.W.F, A.M.F, S.J.S, D.H.H, S.H, M.T, A.F, R.W.T, D.A.S, A.G.C performed functional studies and/or data analysis (including proteomics, immunohistochemistry, molecular biology and cellular experiments including fibroblast culturing, RNA extraction, RNA-sequencing, cDNA synthesis, cloning, Sanger sequencing, Blue Native PAGE and western blots); L.N.S, D.A.S, A.G.C wrote the initial draft of the manuscript. All authors approved the manuscript.

## Notes

### Competing Interest Statement

The authors have declared no competing interest.

### Funding Statement

This study was funded by Australian NHMRC Investigator Fellowships (GNT2009732 DAS, GNT2026315 AF) and a Principal Research Fellowship (GNT1155244 DRT) as well as the Australian Genomics NHMRC Targeted Call for Research grant GNT1113531 and the Australian Medical Research Future Fund Genomics Health Futures Mission (2007959 DRT, 2016030 DAS, 76747 ZS). We thank the Mito Foundation for their support in the form of grants for research (G067 DRT) and provision of equipment (G189 DAS) and a PhD Top-up scholarship (S021 LNS). This work was also supported by grants from The Royal Children’s Hospital (RCH) Foundation (2021-1377). We thank the Bio21 Melbourne Mass Spectrometry and Proteomics Facility (MMSPF) for the provision of instrumentation and training. Work at the MCRI is supported through the Victorian Government’s Operational Infrastructure Support Program. The Chair in Genomic Medicine awarded to JC is generously supported by The RCH Foundation. RWT is funded by the Wellcome Centre for Mitochondrial Research (203105/Z/16/Z), the Medical Research Council (MR/W019027/1), the Lily Foundation, the UK National Institute for Health Research (NIHR) Biomedical Research Centre for Ageing and Age-related disease award to the Newcastle upon Tyne Foundation Hospitals NHS Trust and the UK NHS Highly Specialised Service for Rare Mitochondrial Disorders of Adults and Children. RWT, MT and AMF are funded by LifeArc. CLA is supported by a NIHR Post‐Doctoral Fellowship (PDF‐2018‐11‐ST2‐021), the Lily Foundation and the UK NHS Highly Specialised Service for Rare Mitochondrial Disorders of Adults and Children. Research reported in this publication was supported by the National Institute Of Neurological Disorders And Stroke of the National Institutes of Health (NIH) under Award Number(s) (U01NS134350). This research was supported by JSPS KAKENHI (JP23H00424 and JP23K07236), Japan Agency for Medical Research and Development (AMED) (JP25ek0109672 and JP23ek0109625) and the Ministry of Health, Labour and Welfare of Japan (JP23FC1034). The content is solely the responsibility of the authors and does not necessarily represent the official views of the NHMRC, the NIH, the NHS, the NIHR, or the Department of Health and Social Care.

### Author Declarations

Samples from probands and family members were obtained after receiving written, informed consent for diagnostic or research investigations from the respective responsible human ethics institutional review boards and research was conducted in accordance with the Declaration of Helsinki. Individuals 1 and 2 were consented under HREC/RCH/34228 and HREC/82160/RCHM-2022, approved by the Royal Children’s Hospital, Melbourne (Australia) Ethics in Human Research Committee. Family 3 were enrolled and provided written consent to participate in the Undiagnosed Diseases Network (UDN) NIH (USA) protocol (15-HG-0130) and genomic data was analyzed with an IRB-approved protocol (COMIRB# 16-0146) under the UDN Study protocol approved by the NIH IRB. Individual 4’s parents were consented for genetic testing (trio WES), following NHS (UK) national record of discussion guidelines. Individual 5 was consented under Manchester University NHS (UK) Foundation Trust consent for access to records and stored samples and Genomic Sequencing Consent. Additional ethical approval was granted by the Newcastle and North Tyneside (UK) Research Ethics Committee (REC reference: 16/NE/0267). Individual 6 and the family provided written informed consent under protocols approved by the Saitama Medical University Ethics Review Committee (approval no. 844-VI, Saitama, Japan), and the Medical Research Ethics Committee of Juntendo University (approval no. M17-0089, Tokyo, Japan).

