## Supplemental Files for "*De novo* truncation variants in the low-complexity C-terminal region of *XRN1* are associated with a dominant form of lethal infantile mitochondrial cardiomyopathy"

### SUPPLEMENTAL NOTE: CASE REPORTS

Please contact corresponding author for access to this information.

### SUPPLEMENTAL FIGURES AND LEGENDS

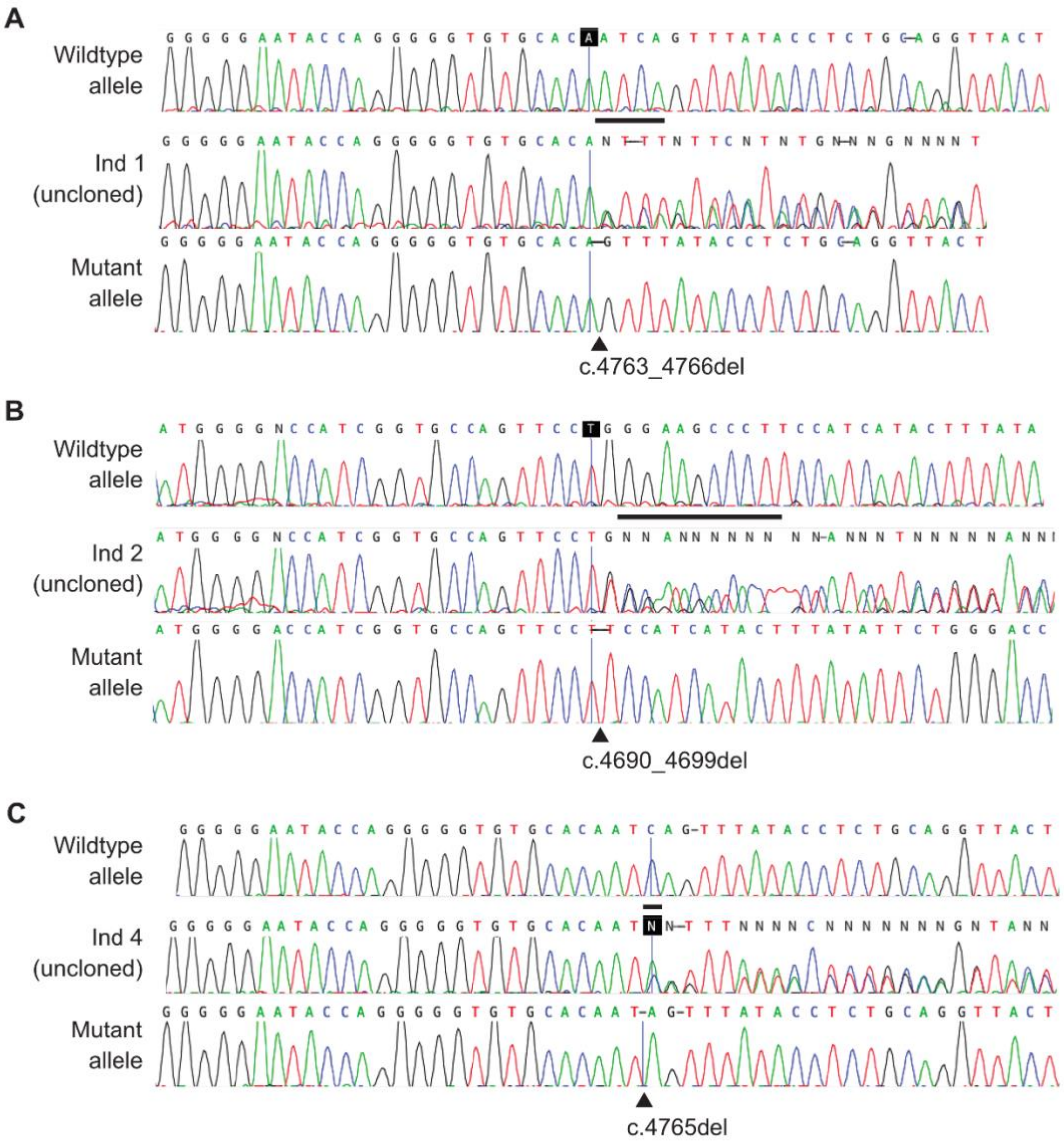


**Figure S1.** ***cDNA sequencing of XRN1 indicates that the mutant alleles do not undergo nonsense-mediated decay.*** Sanger sequencing of *XRN1* from cDNA generated from RNA extracted from Individual 1 and 2 fibroblasts (**A, B**) or Individual 4 skeletal muscle (**C**). Upper and lower images in each panel shows the cloned-out wildtype and mutant alleles, respectively. Middle image is the uncloned product showing the stable expression (escape from nonsense mediated decay) of both wild-type and mutant alleles. Nucleotides deleted by the variant are indicated by a dark bar in the wildtype allele; arrow shows the position of the variant in mutant allele.

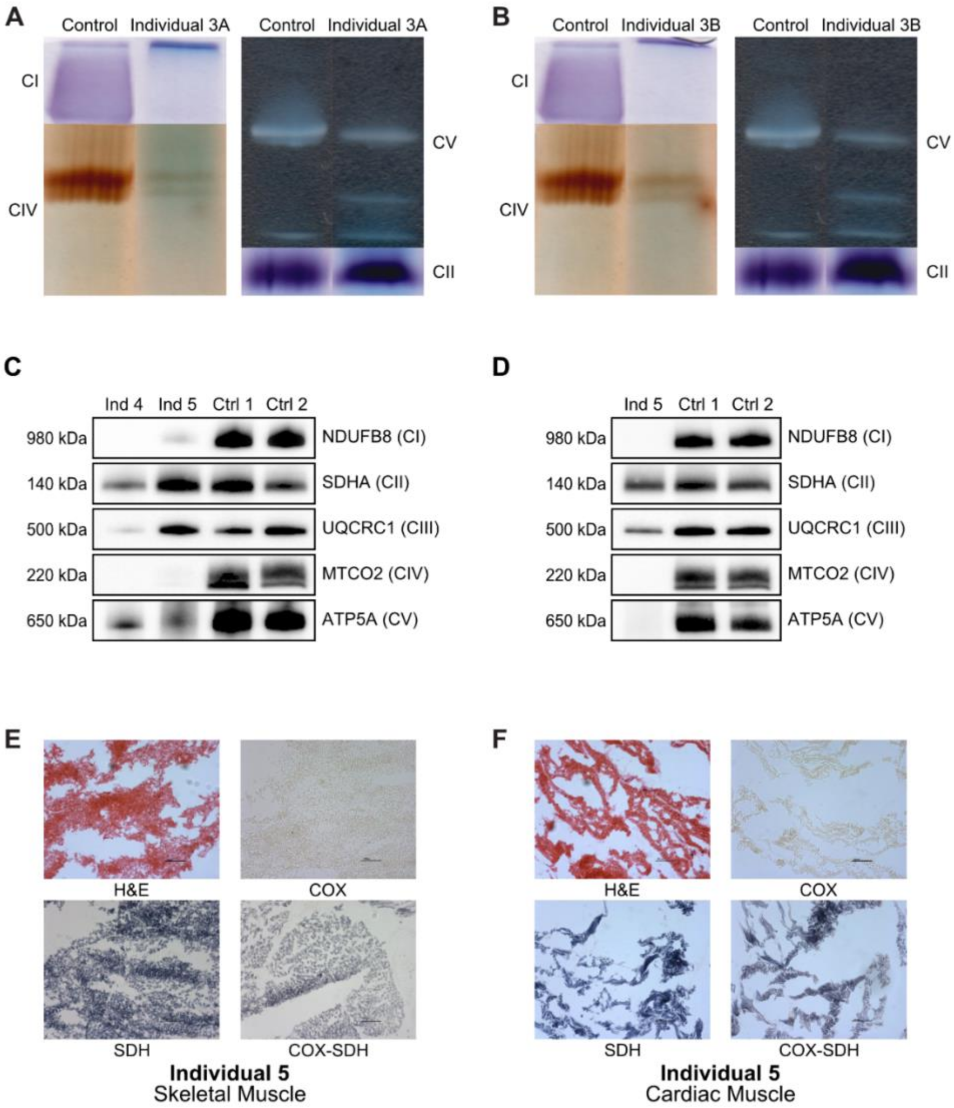


**Figure S2:** ***Blue Native PAGE (BN-PAGE) and immunohistochemistry adds additional support for a combined OXPHOS disorder in striated muscle for Family 3 (Individuals 3A and 3B) and Individuals 4 and 5.*** **(A-B)** Mitochondria isolated from skeletal muscle from Individuals 3A (A) and 3B (B) were separated by BN-PAGE and subjected to an In-Gel Activity assay of individual OXPHOS Complexes I, III, IV and V (CI, CII, CIII and CV). **(C-D)** Mitochondria isolated from skeletal muscle (C) and cardiac muscle (D) of Individual 4 and/or Individual 5 were separated by BN-PAGE and subjected to immunoblotting analysis using antibodies directed to various OXPHOS complexes as indicated. **(E-F)** Histopathological assessment of frozen tissue sections from Individual 5 assessing mitochondrial abnormalities in post-mortem skeletal muscle **(E)** and cardiac muscle (**F**) Hematoxylin and eosin (H&E) staining shows evidence of ice-freezing artefact and poor tissue morphology. Cytochrome *c* oxidase (COX) and sequential succinate dehydrogenase (SDH) histochemistry reveal near global loss of enzyme activities, whilst SDH histochemical activities are preserved. Scale bar = 100 µm.

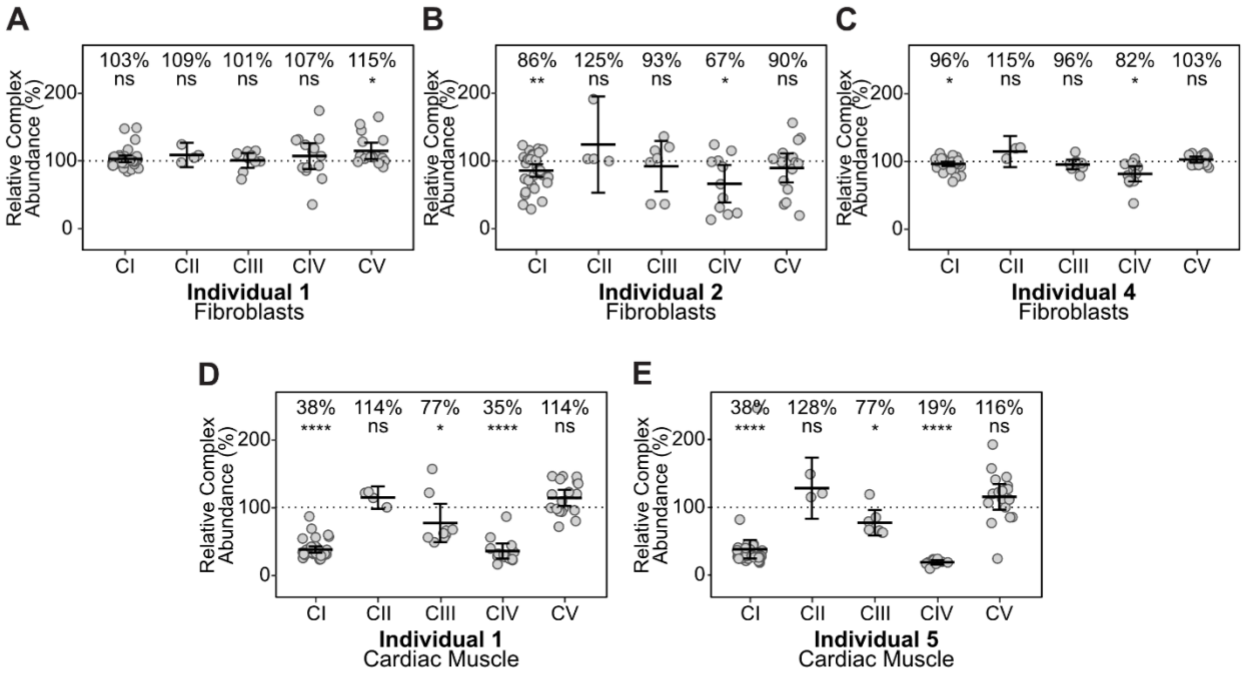


**Figure S3:** ***Quantitative proteomics identifies a combined mitochondrial OXPHOS defects in cardiac tissues for Individual 1 and 5 but not in fibroblasts.*** Relative Complex Abundance (RCA) of OXPHOS Complexes I-V (CI-V) from fibroblast proteomic data (n=5) **(A-C)** and cardiac muscle proteomic data (n=3) **(D-E)** of indicated individual compared to respective controls. Individual protein subunits are represented as a single dot and the mean value of each complex is indicated by the middle bar and represented as a percentage. Upper and lower bars represent 95% confidence intervals. Significance was calculated from a two-sided *t*-test. ns = not significant, * = $p<0.05$, ** = $p\leq0.01$, **** = $p\leq0.0001$

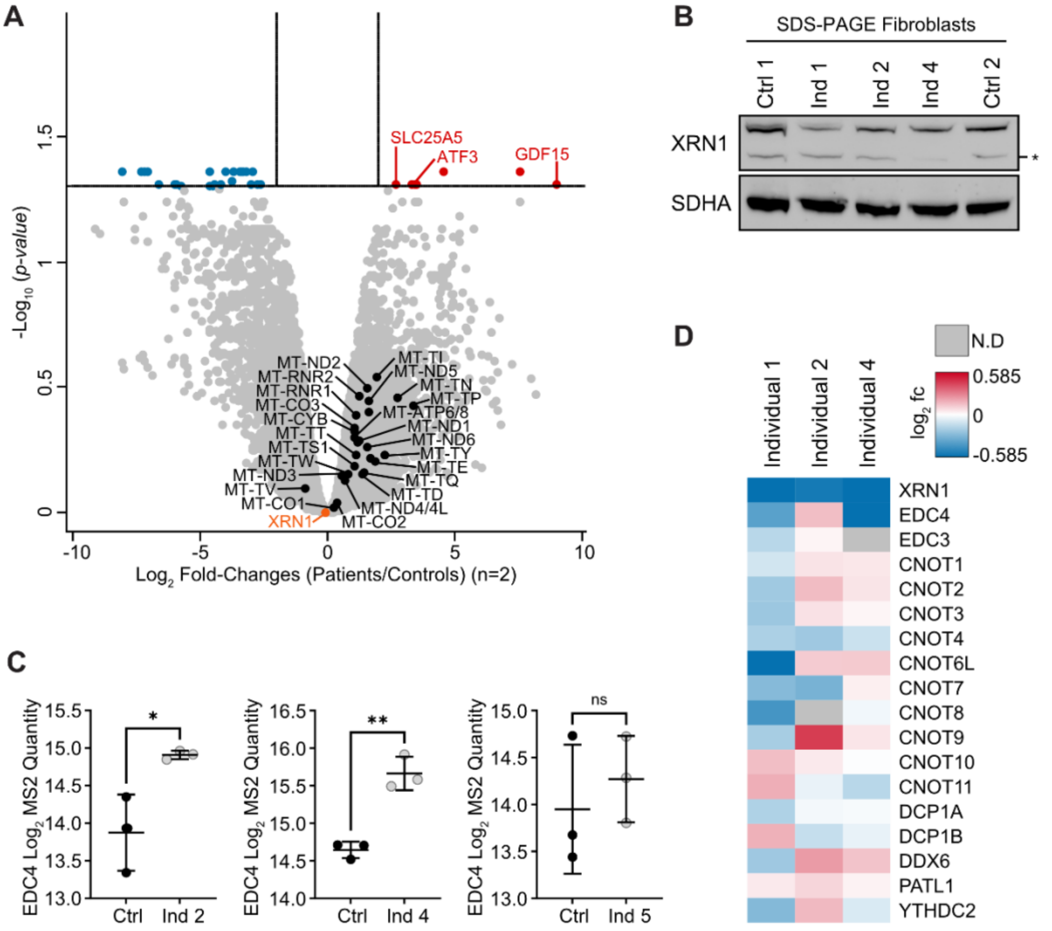


**Figure S4**. ***Functional studies suggest NAD-decapping of mt-RNA is not altered with mt-RNA transcripts unchanged however loss of the EDC4 binding site may alter XRN1:EDC4 ratios.*** **(A)** Volcano plot depicting total RNAseq data showing differentially expressed transcripts detected in Individual 1, 2 and 4 (aggregated data) skeletal muscle samples compared to controls (n=2), demonstrating no significant change in mt-RNA transcripts (Black) along with XRN1 levels (Orange). Vertical lines represent ± 4-fold-change ($\log_2 = \pm 2$) and horizontal line represents *p*-value = 0.05 ($-\log_{10} = 1.301$). Blue = significantly decreased transcripts. Red = significantly increased transcripts. **(B)** Whole cell fibroblast lysates from Individuals 1, 2 and 4 along with two controls were analyzed by SDS-PAGE and immunoblotted against an XRN1 (raised against epitope amino acids 705-785) and loading control (SDHA) antibody as indicated. * = non-specific bands **(C)** Relative abundance of EDC4 protein in skeletal muscle proteomics from Individual 2 (left), Individual 4 (middle) and Individual 5 (right) compared to controls. EDC4 was not detected in Individual 1, 3A or 3B datasets. Significance was calculated from an unpaired t-test between the means. Error bars indicate standard deviation. * = $p \leq 0.05$, ** =

$p \leq 0.01$, ns = not significant. **(D)** Heatmap depicting $\log_2$ fold-changes of 5'-3' mRNA decay proteins in fibroblast proteomics data. Each individual is compared against controls from the same experimental batch. N.D = not detected.

### SUPPLEMANTAL MATERIALS AND METHODS

#### Ethics Statement

Samples from probands and family members were obtained after receiving written, informed consent for diagnostic or research investigations from the respective responsible human ethics institutional review boards and research was conducted in accordance with the Declaration of Helsinki. Individuals 1 and 2 were consented under HREC/RCH/34228 and HREC/82160/RCHM-2022, approved by the Royal Children's Hospital, Melbourne (Australia) Ethics in Human Research Committee. Family 3 were enrolled and provided written consent to participate in the Undiagnosed Diseases Network (UDN) NIH (USA) protocol (15-HG-0130) and genomic data was analyzed with an IRB-approved protocol (COMIRB# 16-0146) under the UDN Study protocol approved by the NIH IRB. Individual 4's parents were consented for genetic testing (trio WES), following NHS (UK) national record of discussion guidelines. Individual 5 was consented under Manchester University NHS (UK) Foundation Trust consent for access to records and stored samples and Genomic Sequencing Consent. Additional ethical approval was granted by the Newcastle and North Tyneside (UK) Research Ethics Committee (REC reference: 16/NE/0267). Individual 6 and the family provided written informed consent under protocols approved by the Saitama Medical University Ethics Review Committee (approval no. 844-VI, Saitama, Japan), and the Medical Research Ethics Committee of Juntendo University (approval no. M17-0089, Tokyo, Japan).

#### Mitochondrial Respiratory Chain Enzyme assays

For Individuals 1 and 2, spectrophotometric enzyme assays assessing mitochondrial respiratory chain and citrate synthase activities in cultured fibroblasts, skeletal muscle, and liver samples were performed as described previously.[5] For Family 3, respiratory chain enzyme activities were determined spectrophotometrically in muscle homogenate as previously described.[6; 7] The results are provided as the activity for Complexes I, II, II+III combined and citrate synthase or as the rate constant for complexes III and IV and compared to the activities in 25 controls. In normal controls, the activity was normally distributed after log transformation, and the patient activities were also expressed as the Z-score. Similarly, the ratio of the activities over the activity of citrate synthase and over the activity of complex II were similarly represented. For Individuals 4 and 5, mitochondrial respiratory chain enzymes were measured spectrophotometrically in cultured fibroblasts and skeletal muscle as previously described.[5; 8] Heart autopsy samples were also tested for Individuals 1 and 5 as per respective enzyme assays above.

**Genomic sequencing and variant detection**

For Individual 1, DNA was extracted from skin fibroblasts and singleton PCR-free genome sequencing (GS) performed (as for P1 and P2 in reference [9]). The mitochondrial genome (mtDNA) along with a targeted analysis of 6843 genes was performed including known mitochondrial disease genes (PanelApp Australia Mitochondrial disease list v0.841), candidate mitochondrial genes predicted or reported to be associated with mitochondrial function[10] and Mendeliome genes described to underlie disease per PanelApp Australia[11] (Mendeliome list v1.418 October 20, 2022). For Individual 2, trio GS and analysis was performed on DNA extracted from blood at a clinically accredited laboratory (Victorian Clinical Genetics Services, Melbourne, Australia), as previously described.[3] Further research GS reanalysis was then performed incorporating the same candidate mitochondrial gene list as Individual 1. When both individuals remained without molecular diagnosis, this research reanalysis was expanded to look at all rare (MAF <0.005), high-impact variants (nonsense, essential splice-site, frameshift type) in all genes regardless of inheritance type. Subsequent segregation studies were performed using PCR on genomic DNA extracted from saliva samples from the parents of Individual 1 using primers: 5’ CCTGAGGTTGAATGTTGTAAGGA 3’ and 5’ TCAGAGAGCAGAGACAGGTCAG 3’ as per [12].

For Family 3, DNA was extracted from whole blood on the trio (Individual 3A, mother and father) with ES with mtDNA sequencing performed through a CAP/CLIA certified laboratory which was non-diagnostic. Through the Undiagnosed Diseases Network, whole blood DNA underwent trio GS in an alternate CAP/CLIA certified laboratory. The initial clinical report was non-diagnostic. Further research re-analysis on the trio ES data and trio GS data was performed utilizing a phenotype agnostic approach to variant prioritization. Sanger confirmation testing was performed on DNA obtained from newborn screening blood spot for the *XRN1* variant identified in the proband (Individual 3A) as well as for segregation studies on their deceased affected twin (Individual 3B) in a CAP/CLIA certified laboratory.

For Individual 4, blood DNA underwent gene agnostic trio ES by the NHS Exeter Genomics Laboratory (R14 test ID, as per NHS Genomic Test Directory https://www.england.nhs.uk/publication/national-genomic-test-directories/). This was initially non-diagnostic, but the data underwent research-based reanalysis in light of the subsequent OXPHOS abnormalities in patient biopsy material with variants prioritized initially based upon having a minor allele frequency <1% and no homozygotes on gnomAD v4. From this rare dataset, variants were stratified according to the anticipated inheritance pattern (recessive, *de novo*, X linked). Variants were retained if predicted to be damaging using the criteria REVEL>0.7 , SpliceAI>0.2 and/or CADD>25. *De novo* variants were prioritized only if they were absent on the gnomAD v4.0 population database, working on the premise that individuals with an acute pediatric disorder would not be represented in the healthy population. X linked variants were prioritized if there were no hemizygote or homozygous individuals on gnomAD.

For Individual 5, GS data was generated by the NHS Genomic Medicine Service and processed using the Genomics England GMS bioinformatics pipeline. Sequence alignment and variant calling were performed using the Illumina DRAGEN Bio-IT Platform (v3.2.22), as described in the Genomics England genomic data source documentation. Initial diagnostic panel-based analysis using Congenica focused on known disease genes and therefore did not initially identify a diagnosis. Following identification of the *XRN1* variant in Individual 4, the NHS Genomic Medicine Service's Clinical Variant Ark was screened for other similar cases. The search focused on rare *de novo* variants situated within the last two exons of *XRN1* and resulted in identification of Individual 5.

For Individual 6 and parents, genomic DNA was extracted from blood using the Maxwell RSC automated extraction platform (Promega). Singleton genome sequencing was performed as per [13]. Variants with minor allele frequencies of <0.5% were filtered from dbSNP, 1KG, the Genome Aggregation Database (gnomAD), and 14KJPN databases of Japanese Multi Omics Reference Panel (jMorp) from the Tohoku Medical Megabank Organization (ToMMo). Manta, Lumpy, ERDS, and CNVnator were used to search for structural variants (SVs) and copy number variants (CNVs). Sanger confirmation in Individual 6's DNA for the *XRN1* variant along with segregation testing of parental genomic DNA was performed using primers: XRN1-4795-F, 5'-ATGCAGGTTGAGCAATCGGA-3', XRN1-4795-R, 5'-GCCACCATGCCTGTCTAGTT-3'.

**mtDNA depletion studies**

The relative abundance of mtDNA vs nuclear DNA was tested in skeletal muscle from Individuals 1 and 2, as well as control skeletal muscle, using quantitative real-time PCR as previously described.[14]

**cDNA studies**

For cDNA studies, RNA was extracted from either cultured fibroblasts or skeletal muscle biopsies from three patients (Individuals 1, 2 and 4) and controls. Cultured fibroblasts were grown in DMEM +10% FBS with and without cycloheximide (100 ng/ml for 24 hours) to inhibit nonsense-mediated decay, as previously described before harvesting.[1]

Total RNA was extracted from fibroblasts using the Illustra RNAspin Mini Isolation Kit (GE Healthcare). While for skeletal muscle, 15 µg of tissue was first sectioned into 8 µm thickness sections using a Leica CM1860 cryostat prior to total RNA extraction using the miRNeasy Mini Kit (Qiagen) per manufacturer's protocols with additional treatment with RNase-Free DNase (Qiagen). cDNA was synthesised using the SuperScript III First-Strand Synthesis System (ThermoFisher Scientific) as per the manufacturer's protocol. The two main *XRN1* transcripts, NM_019001.5 and NM_001282857.2 (MANE transcript), differ only by the inclusion of 3 additional nucleotides at the acceptor-site of exon 36 in NM_001282857.2 and the inclusion of an additional 39bp exon 40 in NM_019001.5. To check the transcript expression and stability of these full-length transcripts, cDNA was amplified using the Expand long range dNTPack kit (Roche) using primers 5' TGGATCTCAGAGCGGTATCC 3'; 5' CAGCACCACAACAAATGTGA 3' were used alongside control PCRs of a similar transcript length (*TOP3A* cDNA 5' CCCGAAGACCGTGCCTTT 3', 5' CTGGCCTGCTCCAGAGTGAT 3' or *SERAC1* 5' GGTCCGACGAGCAGTTGG 3', 5' TGAGCCATTTACCTTCAGATGC 3'). A smaller C-terminal *XRN1* fragment containing exons 39-42 (*XRN1* NM_019001.5) was also amplified for cloning using the pCR2.1-TOPO plasmid, TOPO TA cloning kit (ThermoFisher Scientific) and TOP10 competent cells (ThermoFisher Scientific) as previously described[1] when necessary, using primers 5' TTATCCTTCAGCTGTACCAC 3'; 5' AGAGGACTTCAAAGAAGCTG 3'. Both full-length products and individual cloning colonies were examined on agarose gels and analyzed by Sanger sequencing.

**Quantitative Proteomics**

Skeletal and cardiac muscle from patients and controls were lysed by probe sonication with an amplitude of 30% with 3 pulses of 10 seconds in a lysis buffer containing 5% SDS and 50 mM tetraethylammonium bromide (TEAB) pH 8.2. Samples were then clarified by centrifugation at 6,000 g from 5 minutes. Fibroblast cells were cultured in Dulbecco's Modified Eagle Medium (DMEM) High Glucose, Sodium Pyruvate and Glutamine (Sigma-Aldrich) supplemented with 10% FBS (CellSera), 100 U/mL Penicillin-Streptomycin (Gibco) and 50 µg/mL Uridine. Cells were maintained at 37°C with 5% $CO_2$. Whole cell fibroblast pellets were solubilized the sample lysis buffer and quantified using a Pierce BCA Quantification Kit (ThermoFisher Scientific). For samples analyzed from Individual 1-4, 25 µg of protein was aliquoted in triplicates for patient samples or in single replicates for controls (n=3 for skeletal and cardiac muscle, n=5 for fibroblasts). For samples analyzed from Individual 5, 50 µg of protein was aliquoted in triplicates from patient samples or in single replicates for controls (n=3). All samples were processed  using S-Trap™ columns (Protifi) as previously described.[15; 16] Briefly, solubilized samples were alkylated and reduced with 40 mM chloroacetamide (CAA) and 10 mM Tris(2-carboxyethyl)phosphine (TCEP), followed by overnight digestion with 1 µg of trypsin at 37°C. Eluted peptides were dried down using a CentriVap Concentrator prior to reconstitution in 2% acetonitrile (ACN) and 0.1% trifluoracetic acid (TFA) for analysis by liquid chromatography mass spectrometry (LC-MS/MS).

Tryptic peptides from Individuals 1-4 samples were injected into an Orbitrap Eclipse Mass Spectrometer (ThermoFisher Scientific) equipped with an Acclaim Pepmap nano-trap column (Dinoex-C18, 100 Å, 75 µm x 2 cm) and an Acclaim Pepmap RSLC analytical column (Dionex-C18, 00 Å, 75 µm x 2 cm). The mass spectrometer was operating in a Data Independent Acquisition (DIA) over a 95-minute gradient with a previously described method.[15] Raw files were searched with Spectronaut (v.19.0.240606.62635, Rubin) with an *in silico* library free search for skeletal and cardiac muscle samples using the directDIA+ (deep) workflow, or for fibroblast samples searched against a deeply fractionated data dependent acquired (DDA) spectral library containing 131,627 precursors derived from control fibroblasts. Default BSG factory settings were used with the 'Exclude Single Hits' option selected, and 'Major Top N' along with 'Minor Top N' options were de-selected. UniprotIDs reviewed human canonical and isoforms fasta files (42,386 entries) were used to conduct the search.

Samples analyzed from Individual 5, tryptic peptides were injected into an Evosep One system with a 15 cm Aurora Elite C18 column with integrated captive spray emitter (IonOpticks), at 50 °C. Buffer A was 0.1 % formic acid in HPLC water, buffer B was 0.1 % formic acid in acetonitrile. Immediately prior to LC-MS, peptides were resuspended in buffer A and a volume equivalent to 500 ng was loaded onto the LC system-specific C18 EvoTips, according to manufacturer instructions, and subjected to the predefined Whisper-Zoom 20 SPD (where the gradient is 0-35 % buffer B, 200 nl/min, for 58 minutes, 20 samples per day were permitted) using the Evosep One (Evosep) in line with a timsToF HT mass spectrometer (Bruker) operated in diaPASEF mode. Mass and IM ranges were 300-1200 *m/z* and 0.6-1.45 $1/K_0$, diaPASEF was performed as previously described[17] using variable width IM-*m/z* windows without overlap. TIMS ramp and accumulation times were 100 ms, total cycle time was ~1.8 seconds. Collision energy was applied in a linear fashion, where ion mobility = 0.6-1.6 $1/K_0$, and collision energy = 20 - 59 eV. Raw diaPASEF data files were searched using DIA-NN V 2.0.1[18], using its *in silico* generated spectral library function, based on reference proteome FASTA files for *H. sapiens* (UP000005640, SwissProt with isoforms, downloaded from UniProt on 11/05/2023) and a common contaminants list.[19] Trypsin specificity with a maximum of 2 missed cleavages was permitted per peptide, cysteine carbamidomethylation were set as a fixed modification, oxidation of methionine and protein N-terminal acetylation as variable (with up to two variable modifications per peptide). Peptide length and m/z were 7-30 and 300-1200, charge states 2-4 were included. Mass accuracy was fixed to 15 ppm for MS1 and MS2. Protein and peptide FDR were both set to 1 %. All other settings were left as default.

All raw proteomic data across Individuals 1-5 was processed using Perseus (1.6.15.0) platform.[20] Raw MS2 values were $\log_2$ transformed and filtered for at least 2 values in each experimental group. A two-sided *t*-test was carried out on a whole-cell and mitochondrial proteome level. For the mitochondrial proteome, mitochondrial proteins were annotated by MitoCarta3.0 and subsetted before being normalized by subtracting the mean of all datapoints in individual sample replicates to account for differences in mitochondrial content. Normalized values were then subjected to a student *t*-test. Heatmaps were generated by exporting $\text{Log}_2$ fold-changes for from either whole-cell or mitochondrial normalized *t*-tests and visualized by heatmap using Morpheus (https://software.broadinstitute.org/morpheus/). Relative Complex Abundance analysis was carried out on raw MS2 quantities of mitochondrial proteins using an in-house *R* script as previously described.[15]

Enrichment Analysis was performed on proteins significantly changed in skeletal muscle proteomics data ($p$-value ≤ 0.05, fold-change ± 1.5) compared to controls, from at least two of the following Individuals; Individual 1, 2, 4 and 5. Individuals 3A and 3B datasets were excluded from this analysis as overall detection of non-mitochondrial proteins were lower than that of the other datasets. Inclusion of mitochondrial proteins were based on $p$-value and fold-change values of normalised mitochondrial proteomic $t$-test, to account for changes in total mitochondrial content between affected individuals and controls. Significantly increased and decreased proteins were analyzed separately. The clusterProfiler[21] package in *R* was used to perform the over-representation analysis using WikiPathways annotations, against a background list of all proteins detected in skeletal muscle datasets across the 4 individuals. Pathways with adjusted $p$-value <0.05 were considered significantly enriched. Results were visualized in R with dot size representing the number of genes detected for that pathway and color scale indicating adjusted $p$-value.

**SDS-PAGE and Immunoblotting**

Whole cell lysates were prepared from cultured fibroblasts grown in DMEM + 10% FBS for Individuals 1, 2, 4 and controls as previously described[9] using extraction buffer containing 1.5% n-Dodecyl-β-d-maltopyranoside, 25 mM HEPES and 100 mM NaCl) with further solubilization in sodium dodecyl sulfate (SDS)/glycerol solubilization buffer (125mM Tris pH8.8, 40% glycerol, 4% SDS, 100mM DTT, 0.01% Bromophenol blue, protease inhibitor cocktail) after protein concentrations were determined using BCA analysis,[22] and analysis by SDS-PAGE as described previously.[1] SDS-PAGE gels were transferred onto polyvinylidene difluoride (PVDF) membrane and probed with primary antibodies raised against XRN1 (1:500; ThermoFisher Scientific; PA5-57110), and SDHA (1: 10,000; Invitrogen; 459200) as a loading control for mitochondrial protein content. Blots were incubated with anti-mouse or anti-rabbit IgG secondary antibodies (Cytiva, mouse #GEHENA931 and rabbit #GEHENA934) and developed with Clarity Western ECL Substrate (Bio-Rad Laboratories) and visualized using the ChemiDoc Imaging System (Bio-Rad Laboratories).

**Blue Native PAGE and Immunoblotting**

For Individuals 3A and 3B skeletal muscle samples, mitochondria were prepared by differential centrifugation, after solubilization of the inner mitochondrial membrane fraction were separated on a non-denaturing blue native polyacrylamide gel electrophoresis (BN-PAGE) and the in-gel activities of complexes I, II, IV and V determined by the formation of colored precipitate as previously described.[6; 7; 23] For Individuals 4 and 5 skeletal and/or cardiac muscle, mitochondrial fractions were solubilized separated on a non-denaturing BN-PAGE gel, as above. BN-PAGE gels were transferred onto PVDF membrane and probed with primary antibodies raised against NDUFB8 (complex I) (1:1000; Abcam, 110242), SDHA (complex II) (1:1000; Abcam, 14715), UQCRC2 (complex III) (1:1000; Abcam, 14745), COXI (complex IV) (1:1000; Abcam, 14705) and ATP5A (complex V) (1:1000; Abcam, 14748). Blots were incubated with anti-mouse IgG secondary antibodies (1:2000; Agilent Technologies, P026002-2) and developed with SuperSignal West Pico Plus (Life Technologies) and visualized using the ChemiDoc Imaging System (Bio-Rad Laboratories).

**Histopathological analyses**

10 μM of frozen skeletal and cardiac muscle sections were used in each assessment. For histopathological studies, H&E staining was employed to determine muscle morphology, whereas sequential COX and SDH histochemistry was used to assess COX activity in muscle fibers. Quadruple immunofluorescence assays were carried out by measuring NDUFB8 (CI) and MT-CO1[24] protein abundance against the mitochondrial mass marker, porin, using in-house analysis software as previously outlined.[25]

**RNA sequencing for mt-RNA transcripts**

To assess mitochondrial RNA transcript abundance and processing, RNA was extracted from Individual 1, 2 and 4, as well as control skeletal muscles using the miRNeasy Mini Kit (Qiagen), and RNA was treated with RNase-Free DNase (Qiagen). RNA quality and quantity were tested using TapeStation RNA ScreenTape analysis (Agilent) and Qubit RNA HS (ThermoFisher), respectively. RNA samples had an RNA integrity number of 6.5–8.3. The Illumina stranded total RNA Prep Kit with Ribo-Zero plus reagent (Illumina) was used to generate sequencing libraries from 100ng RNA and paired-end sequencing on a NovaSeq 6000 (Illumina) was performed to achieve ~80 million read coverage per sample. RNA-seq data was aligned to the human genome (GRCh38) with STAR 2.7.10b[26] in a three-pass process that included a third mitochondrial realignment step to prevent the spurious introduction splice junctions to the mitochondrial genome (--alignIntronMax 1). Exon-spanning reads were counted with featureCounts 2.0.6[27] (--countReadPairs -p -B -Q 255 -s 2) and differential gene expression of patients against controls was analyzed with limma 3.30.6[28] and voom using the GENCODE v41 basic release with a custom mitochondrial annotation, collapsed to meta-genes, and masked for NuMTs sequences. Genes with a log fold change greater or less than $\log_2(1.2)$ and an adjusted p-value of less than 0.05 were considered significant. RNA-seq data from single biological replicates of Individuals 1, 2, and 4 were aggregated and treated as a single condition 'patients' for differential expression analysis against controls (n=2). A volcano plot was visualized using the scatterplot function in Perseus (1.6.15.)[20], plotting the $\log_2$ fold-change against $-\log_{10}$ transformed adjusted *p*-values.

***ACKNOWLEDGMENTS***

We thank Dr Langping He (Newcastle upon Tyne Hospitals NHS Foundation Trust) and Dr Amanda Lam (Institute of Neurology, Queen Square, London) for their help with the assessment of mitochondrial respiratory chain activities. This research was supported by Australian NHMRC Investigator Fellowships (GNT2009732 DAS, GNT2026315 AF) and a Principal Research Fellowship (GNT1155244 DRT) as well as the Australian Genomics NHMRC Targeted Call for Research grant GNT1113531 and the Australian Medical Research Future Fund Genomics Health Futures Mission (2007959 DRT, 2016030 DAS, 76747 ZS). We thank the Mito Foundation for their support in the form of grants for research (G067 DRT) and provision of equipment (G189 DAS) and a PhD Top-up scholarship (S021 LNS). This work was also supported by grants from The Royal Children's Hospital (RCH) Foundation (2021-1377). We thank the Bio21 Melbourne Mass Spectrometry and Proteomics Facility (MMSPF) for the provision of instrumentation and training. Work at the MCRI is supported through the Victorian Government's Operational Infrastructure Support Program. The Chair in Genomic Medicine awarded to JC is generously supported by The RCH Foundation. RWT is funded by the Wellcome Centre for Mitochondrial Research (203105/Z/16/Z), the Medical Research Council (MR/W019027/1), the Lily Foundation, the UK National Institute for Health Research (NIHR) Biomedical Research Centre for Ageing and Age-related disease award to the Newcastle upon Tyne Foundation Hospitals NHS Trust and the UK NHS Highly Specialised Service for Rare Mitochondrial Disorders of Adults and Children. RWT, MT and AMF are funded by LifeArc. CLA is supported by a NIHR Post-Doctoral Fellowship (PDF-2018-11-ST2-021), the Lily Foundation and the UK NHS Highly Specialised Service for Rare Mitochondrial Disorders of Adults and Children. Research reported in this publication was supported by the National Institute Of Neurological Disorders And Stroke of the

National Institutes of Health (NIH) under Award Number(s) (U01NS134350). This research was supported by JSPS KAKENHI (JP23H00424 and JP23K07236), Japan Agency for Medical Research and Development (AMED) (JP25ek0109672 and JP23ek0109625) and the Ministry of Health, Labour and Welfare of Japan (JP23FC1034). The content is solely the responsibility of the authors and does not necessarily represent the official views of the NHMRC, the NIH, the NHS, the NIHR, or the Department of Health and Social Care.

**UNDIAGNOSED DISEASES NETWORK**

Aaron Quinlan, Abdul Elkadri, Alan H. Beggs, Albert R. La Spada, Alden Huang, Aleksandra Foksinska, Alex Paul, Alistair Ward, Alyson Krokosky, Alyssa A. Tran, Andrea Gropman, Andres Vargas, Andrew B. Crouse, Andrew Stergachis, Anna Hurst, Anna Raper, Anne Slavotinek, Arian Nouraee, Arjun Tarakad, Ashley Andrews, Ashley McMinn, Ayuko Iverson, Barbara N. Pusey Swerdzewski, Ben Afzali, Ben Solomon, Beth A. Martin, Brandon M Wilk, Breanna Mitchell, Brendan C. Lanpher, Brendan H. Lee, Brent L. Fogel, Brett Bordini, Brett H. Graham, Bruce Gelb, Bruce R Korf, Camilo Toro, Cara Skraban, Carlos A. Bacino, Carlos A. Pardo-Villamizar, Carlos Prada, Carson A. Smith, Cathy Shyr, Cecilia Esteves, Changrui Xiao, Charlotte Cunningham-Rundles, Chloe M. Reuter, Christine M. Eng, Christopher Mayhew, Chun-Hung Chan, Colleen E. Wahl, Corrine K. Welt, Cynthia J. Tifft, D Isum Ward, Dana Kiley, Dana Sayer, Daniel J. Rader, Daniel Wegner, Danny E. Miller, Daryl A. Scott, Dave Viskochil, David A. Sweetser, David Chiang, David R. Adams, Deborah Barbouth, Deepak A. Rao, Devin Oglesbee, Devon Bonner, Donald Basel, Donna Novacic, Dustin Baldridge, Elaine Seto, Elisabeth Rosenthal, Elizabeth A Worthey, Elizabeth A. Burke, Elizabeth Blue, Elizabeth C. Chao, Elizabeth Wohler, Ellen F. Macnamara, Elsa Balton, Emily Glanton, Emily Shelkowitz, Eneida Mendonca, Eric Allenspach, Eric Gamazon, Eric Gayle, Eric Klee, Eric Vilain, Erica Davis, Erin Conboy, Erin E. Baldwin, Esteban C. Dell'Angelica, Euan A. Ashley, F. Sessions Cole, Filippo Pinto e Vairo, Frances High, Francesco Vetrini, Francis Rossignol, Francisco Bustos, Fuki M. Hisama, Gabor Marth,Gail P. Jarvik, Ganesh Mochida, George Carvalho, Gerard T. Berry, Ghayda Mirzaa, Giorgio Sirugo, Gonench Kilich, Guney Bademci, Hector

Rodrigo Mendez, Heidi Wood, Herman Taylor, Holly K. Tabor, Hongzheng Dai, Hsiao-Tuan Chao, Hugo J. Bellen, Ian Glass, Ian R. Lanza, Ingrid A. Holm, Isaac S. Kohane, Ivan Chinn, J. Carl Pallais, Jacinda B. Sampson, James P. Orengo, James Verbsky, Jared Sninsky, Jason Hom, Jason Schend, Jennefer N. Kohler, Jennifer Morgan, Jennifer Schymick, Jennifer Tousseau, Jennifer Wambach, Jiayu Fu, Jill A. Rosenfeld, Jimann Shin, Joanna Jen, Joanna M. Gonzalez, John A. Phillips III, John Carey, John E. Gorzynski, Joie Davis, Jonathan A. Bernstein, Jose Abdenur, Joseph Loscalzo, Joy D. Cogan, Julian A. Martínez-Agosto, Julie Hoover-Fong, Julie McCarrier, Kahlen Darr, Kai Lee Yap, Kaitlin Callaway, Kathleen A. Leppig, Kathleen A. Sisco, Kathleen Page, Kathleen Sullivan, Katrina Dipple, Kayla M. Treat, Kelly Regan-Fendt, Kelly Schoch, Kevin S. Smith, Khurram Liaqat, Kim Worley, Kimberly Ezell, Kimberly LeBlanc, Kirsten Blanco, Kumarie Latchman, Lakshitha Perera, Lance H. Rodan, Laura Keehan, Lauren Blieden, Lauren C. Briere, Lauren Jeffries , Laurens Wiel, Layal F. Abi Farraj, Leoyklang Petcharet, LéShon Peart, Lili Mantcheva, Lilianna Solnica-Krezel, Lindsay C. Burrage, Lindsay Mulvihill, Lisa Bastarache, Lisa Schimmenti, Lorenzo Botto, Lorraine Potocki, Louise Bier, Lynette Rives, Lynne A. Wolfe, Mafalda Barbosa, Maija-Rikka Steenari, Manish J. Butte, Manisha Balwani, Margaret Delgado, María José Ortuño Romero , María Paula Silva, Maria T. Acosta, Marie Morimoto, Mariya Shadrina, Mark Wener, Marla Sabaii, Martha Horike-Pyne, Martin G. Martin, Martin Rodriguez, Mary Koziura, Matt Velinder, Matthew Might, Matthew Robinson, Matthew T. Wheeler, May Christine V. Malicdan, Megan Bell, Meghan C. Halley, Melissa Walker, Mia Levanto, Michael Bamshad, Michael F. Wangler, Michael Muriello, Michael T. Zimmermann, Miranda Leitheiser, Mohamad Mikati, Mohamad Saifeddine, Monika Weisz Hubshman, Monte Westerfield, Mustafa Tekin, Naghmeh Dorrani, Nara Sobreira, Neil H. Parker, Neil Hanchard, Nicholas Borja, Nicola Longo, Nicole M. Walley, Odelya Kaufman , Orpa Jean-Marie, Page C. Goddard, Paolo Moretti, Patricia Dickson, Patrick McMullen, Paul Auwaerter, Paul Berger, Paul G. Fisher, Pengfei Liu, Peter Byers, Philip Dane Witmer, Pinar Bayrak-Toydemir, Pongtawat Lertwilaiwittaya, Precilla D'Souza, Queenie Tan, Rachel A. Ungar, Rachel Evard, Rachel Li, Rakale C. Quarells, Ramakrishnan Rajagopalan, Raquel L. Alvarez, Reaford Blackburn, Rebecca C. Spillmann, Rebecca Ganetzky, Rebecca Overbury, Rebekah Barrick, Richard A. Lewis, Richard Chang, Rizwan Hamid, Rong Mao, Ronit Marom, Rosario I. Corona, Runjun Kumar, Russell Butterfield, Sanaz Attaripour, Sandesh Nagamani, Saskia Shuman, Seema R. Lalani, Seth Perlman, Shamika Ketkar, Shilpa N. Kobren, Shinya

Yamamoto, Shruti Marwaha, Sirisak Chanprasert, Stanley F. Nelson, Stephan Zuchner, Stephanie Bivona, Stephanie M. Ware, Stephen B Montgomery, Stephen C. Pak, Steven Boyden, Suha Bachir, Susan Shin, Tahseen Mozaffar, Tanner D Jensen, Taylor Beagle, Taylor Maurer, Teneasha Washington, Teodoro Jerves Serrano, Terra R. Coakley, Thomas Cassini, Thomas J. Nicholas, Timothy Schedl, Tina Truong, Tiphanie P. Vogel, Vaidehi Jobanputra, Valerie V. Maduro, Vandana Shashi, Virginia Sybert, Wendy Chung, Wendy Introne, Wendy Raskind, Willa Thorson, William A. Gahl, William E. Byrd, William J. Craigen, Winston Timp, Yan Huang, Yigit Karasozen, Yong-Hui Jiang , Yuka Manabe, Zackary Dov Berger, Ziyuan Guo
